# Mental disorders and discrimination: a prospective cohort study of young twin pairs in Germany

**DOI:** 10.1101/2023.11.01.23297942

**Authors:** Lucas Calais-Ferreira, Greg Armstrong, Elisabeth Hahn, Giles Newton-Howes, James Foulds, John L Hopper, Frank M Spinath, Paul Kurdyak, Jesse T Young

## Abstract

**Background:** Mental disorders and discrimination share common risk factors. The association between having a mental disorder and experiencing discrimination is well-known, but the extent to which familial factors, such as genetic and shared environmental factors, might confound this association, including gender differences in familial confounding, remains unexplored.

**Aims:** We investigated potential unmeasured familial confounding in the relationship between mental disorders and discrimination.

**Method:** We examined 2,044 same-sex twin pairs aged 16–25 years from the German population-based study *’TwinLife’*. We used a matched design and random-effects regression applied to within-individual and within-and-between pair models for the association between mental disorder and discrimination, and used likelihood ratio tests (LRTs) to compare these models. Multivariable models were adjusted for body-mass-index, educational attainment, and global life satisfaction.

**Results:** Mental disorder and discrimination were associated in the adjusted within-individual model (adjusted odds ratio=2.19, 95% Confidence Interval:1.42–3.39, *P*<0.001). However, the within-and-between pair model showed that this association was explained by the within-pair mean (aOR=4.24, 95%CI:2.17–8.29, *P*<0.001) and not the within-pair difference (aOR=1.26, 95%CI:0.70–2.28, *P*=0.4) of mental disorder. Therefore, this association was mostly explained by familial confounding, which is also supported by the LRTs for the unadjusted and adjusted models (*P*<0.001 and *P*=0.03, respectively). This familial confounding was more prominent for males than females.

**Conclusions:** Our findings show that the association between mental disorder and discrimination is almost fully explained by unmeasured familial factors. Incorporating family members in interventions targeted at ameliorating mental ill-health and experiences of discrimination among adolescents may improve efficacy.

## INTRODUCTION

Mental disorders and the experience of discrimination are commonplace in our society, and they are part of a cycle that keeps large portions of our society in pockets of disadvantage and exclusion.^1, 2^ It is estimated that nearly one in three adolescents meet established lifetime criteria for mental disorders,^3^ and mental health problems in 7-17 year-olds have substantially increased as a result of the COVID-19 pandemic.^4^ The prevalence of self-reported discrimination (for the last five years) based on ethnic or immigrant background, skin colour, or religion in German adults is nearly 40%.^5^ Discrimination has been linked to multiple negative social and health outcomes,^6^ and the risk of experiencing discrimination can reach 80% for young people (15-25 years) facing multiple sources of social disadvantage.^7^

Gender is also a key aspect of discrimination, including its intersection with ethnicity, which may start as early as the transition from childhood to adolescence.^8^ Discrimination faced by young people is associated with social determinants of health such as education,^9^ family income,^10^ and ethnic background,^11, 12^ that commonly cluster among families. This familial clustering occurs largely because most people are likely to have the same or very similar socioeconomic characteristics to their immediate (first-degree) family members from a young age.

### Familial factors in mental disorders and discrimination

Through studying twins and other sets of family members, it is now largely understood that individual differences in mental disorders are also likely to be caused, at least partially, by a combination of familial factors of genetic and environmental (including epigenetic) nature.^13^ Families of people with a mental disorder are more likely to experience social isolation and economic hardship.^14^ An earlier onset of mental ill-health has been associated with decreased levels of treatment access and poorer health and social outcomes compared to those who develop symptoms later in life.^15^

A potentially bidirectional and iterative association between mental health and discrimination has been suggested in the literature.^1^ For example, people who reported being discriminated against are at higher risk of being diagnosed with a generalised anxiety disorder (GAD),^16^ depression, and suicidality.^17^ People with mental disorders are more likely to have a future experience of discrimination, in part due to the stigma associated with mental disorders.^18^ Hypotheses about whether the onset of mental disorders produces an effect on the likelihood of discrimination or if the experience of discrimination precedes the onset of serious mental disorders^19^ remain an unsettled debate in psychiatric research.

The extent to which associations between mental disorders and discrimination are confounded by familial factors is largely unknown. Understanding the nature of this relationship is critical because if this association is affected by familial confounding, youth mental health and/or discrimination interventions targeted at the individual level might not produce the desired clinical effect. It is possible to adjust for familial confounding in a given association by studying twin pairs who are, by design, uniquely matched for age, potentially sex, and several familial factors such as their shared home environment, household socioeconomic status, and (at least partially) their genetic factors.^20^

Therefore, using a matched co-twin paired design in a large cohort of adolescent and young adult twins of both sexes in Germany, we aimed to: (1) investigate the within-individual association between any previously diagnosed mental disorder and perceived discrimination in both unadjusted and adjusted models; (2) determine if the association between mental disorder and perceived discrimination are confounded by familial factors; and (3) determine if familial confounding differs by sex.

## METHODS

This was a prospective study following two cohorts of young (16 and 25 years of age on average at baseline) same-sex twin pairs from the TwinLife Study.^21, 22^ The TwinLife Study has recruited and collected comprehensive data from twin pairs and their family members in Germany through a combination of in-person visits to families every two years and phone interviews in the intervening years. Our study used data from the first wave (e.g., baseline) conducted in 2014/2015 and the second wave (e.g., follow-up) in 2016/2017, both through face-to-face household visits. A total of 4,088 twins from 2,044 same-sex twin pairs were included in our study.

### Outcome

Our outcome of interest was defined as a recent experience of discrimination obtained through self-report to the question *“During the last 12 months, have you felt that you were disadvantaged or discriminated against due to any personal characteristics (e.g., your ethnic or cultural background, gender, religious beliefs)?”.* We considered a participant’s report of experiencing discrimination in either waves 1 or 2 as positive for discrimination.

### Baseline measures

Our primary exposure, a previous diagnosis of a mental disorder, was ascertained through self-report in the baseline survey (wave 1) through the question *“Has a doctor ever diagnosed you with one or more of the following illnesses?”.* Participants who responded yes to anxiety disorder, depression, alcohol use disorder, attention-deficit hyperactivity disorder (ADHD) or sleep disorder were considered a positive for mental disorder (dichotomised as a yes/no variable).

Sex was ascertained from the original recruitment process based on community registration offices and was confirmed at the first home visit. Only same-twin pairs were originally recruited for the Twinlife Study. Other measures ascertained at baseline included body mass index (BMI), current smoker status (yes/no), and global life satisfaction measured as a compiled score (from 5-25) from five different domains. This global life satisfaction measure included participant ratings (from strongly disagree to strongly agree on a 5-point scale) for the following statements: *“In most ways, my life is close to my ideal”, “the conditions of my life are excellent”, “I am satisfied with my life”, “so far I have gotten the important things I want in life”, and “if I could live my life over, I would change almost nothing”*.^23^

Self-report of whether the participant had left school before obtaining a primary or secondary school completion certificate (yes/no) was ascertained from both baseline and follow-up surveys (i.e., wave 1 and 2) such that a positive response in either survey was considered positive for leaving school prematurely. Migrant status was ascertained through the self-reported country of origin. Those who reported being from Germany were coded as 0, and twin pairs where at least one twin was recorded as being from another country were coded as 1. Age was matched between the twin pairs and was ascertained at baseline.

### Statistical analyses

Descriptive statistics were calculated for all measures. We used Chi-squared tests and non-parametric Wilcoxon rank-sum tests to assess differences between groups for binary and continuous variables, respectively. We calculated sex-adjusted intra-class correlations (ICC) within twin pairs for all covariates and a sex-adjusted familial risk ratio (FRR)^24^ for mental disorder.

In our regression analysis, which included MZ and DZ pairs together, we first fitted the within-individual models to study the univariable and multivariable associations between risk factors with discrimination. Second, we fitted the within-and-between pair models to study the same associations, with the addition of adjusting for unmeasured familial confounding, such as genetic and shared environmental factors. This approach has been used to investigate if associations between exposures and outcomes are, at least partially, due to unmeasured familial confounders shared by twin pairs.^25, 26^

To study the within-individual association of mental disorder and discrimination (aim 1), we fitted univariable and multivariable logistic regression models with random effects applying maximum likelihood estimation of odds ratios (OR). This allowed us to study within-individual associations between exposure and outcome (including covariates) while accounting for the paired structure of the data to make inferences about individual differences. Our multivariable models were adjusted for sex and for other risk factors with previous evidence of an association with mental health and discrimination: obesity,^27, 28^ educational attainment,^29, 30^ and life satisfaction.^10, 31^ Migrant status, which has also been linked to mental health and discrimination^8, 32^ was a shared variable for all twin pairs. Therefore, regression models including migrant status were only included in the sensitivity analysis (Supplementary Material).

To investigate the presence of, and adjust for, familial confounding in the studied association (aim 2), we fitted within-and-between pair models, also using random effects and maximum likelihood estimation. This approach fits the within-pair difference (the difference between the individual’s value and the within-pair mean) and the within-pair mean separately for each risk factor in the model, allowing disaggregation of their shared (familial) and non-shared contributions to variance in discrimination. When examined using a likelihood ratio test (LRT), if the within-pair difference and the within-pair mean differ statistically, there is evidence that the association is confounded by unmeasured factors, presumed to be familial.

For such within-and-between pair models, the within-pair difference association with the outcome is a measure of the association unconfounded by familial factors, and therefore, the inference is made about related people or people of similar familial risks. In contrast, the between-pair estimate is a measure of the association confounded by unmeasured familial factors. This simulates inference about individuals of ’unknown’ familial similarity, such as in the general population.

More specifically, for binary covariates, the within-pair difference OR indicates the change in risk of the outcome for those with the risk factor compared to their co-twin unaffected by such risk factor. The between-pair difference odds ratio describes the change in risk for twins in pairs where both have the risk factor compared to twins from pairs with no twins affected by that risk factor. If the OR for the within-pair difference attenuates to the null, then this is consistent with the association being confounded (partially or entirely) by familial confounding.

We tested for interactions between the between-pair difference of each covariate and sex in separate models to obtain evidence of sex differences in familial confounding for each of these risk factors (aim 3). We also present our regression analyses stratified by sex in the Supplementary Material.

We conducted sensitivity analyses, first restricting the regression analyses to those who experienced discrimination at wave 2 only (Supplementary Material, Table S1) and to those who experienced in both waves 1 and 2 (Table S2). This was done to ensure a longitudinal relationship between mental disorder and discrimination and to assess the precision of our primary outcome, respectively. We also included unadjusted and adjusted models with migrant status as a risk factor (Table S3) to assess the univariable association between migrant status and discrimination and to test if including the migrant variable in the adjusted model would produce substantial changes in the other estimates.

We fitted within-and-between models separately for male and female twin pairs and included information on interactive terms between sex and the between-pair difference of each covariate to assess sex differences in familial confounding (Table S4). We fitted a model restricting our definition of mental disorder to a diagnosis of anxiety or depression (Table S5) instead of the more heterogeneous mental disorder variable.

We conducted ’complete case’ analyses for both within-individual and within-and-between pair models, whereby any individual or twin pair with missing data were excluded from the analysis. All analyses were conducted in Stata MP 16.0.^33^ This study was reported according to the STROBE statement.^34^

### Ethics statement

The authors assert that all procedures contributing to this work comply with the ethical standards of the relevant national and institutional committees on human experimentation and with the Helsinki Declaration of 1975, as revised in 2008. All procedures involving human subjects/patients were approved by the University of Melbourne’s Human Research Ethics Committee (HREC ref#:21305). Ethics approval for the original TwinLife study has been obtained through the German Psychological Association (protocol numbers: RR 11.2009 and RR 09.2013). Written or verbal informed consent was obtained from all study participants.

## RESULTS

### Descriptive statistics

Four participants were excluded for not having zygosity information. There were 1,022 monozygotic (MZ) and 1,020 dizygotic (DZ) same-sex pairs included in the analysis. Missing data was present for BMI (11%), current smoker status (1%), and global life satisfaction score (2%).

Table 1 provides descriptive statistics of the study sample. The mean age at baseline was 19.9 (Interquartile range = 17–23) years. There were 604 (14.8%) participants who self-reported discrimination at any wave. Around 8% (n=332) of the sample reported having been previously diagnosed with a mental disorder at baseline, and no twins reported a new diagnosis between waves 1 and 2. Approximately 5% of the sample (n=186) were migrants. Of those, 57 (31%) were recorded as being born in a country of the former Soviet Union, 28 (15%) from Eastern Europe and 26 (14%) from Arabic-Islamic countries.

**Table 1:** Descriptive characteristics of study sample.

| Risk factors<sup>1</sup> | Self-reported discrimination (n=604) | No self-reported discrimination (n=3480) | All (n=4084) | *p*<sup>2</sup> |
| --- | --- | --- | --- | --- |
| Diagnosed mental disorder, n (%) | 90/604 (14.9) | 242/3480 (7.0) | 332/4084 (8.1) | <0.001 |
| Male sex, n (%) | 217/604 (35.9) | 1511/3480 (43.4) | 1728/4084 (42.3) | 0.001 |
| Age, years, mean (SD) | 20.0 (3.1) | 19.9 (3.1) | 19.9 (3.1) | 0.2 |
| BMI, kg, mean (SD) | 22.5 (4.3) | 22.2 (3.9) | 22.2 (3.9) | 0.2 |
| Current smoker, n (%) | 150/601 (25.0) | 840/3445 (24.4) | 990/4046 (24.5) | 0.8 |
| Left school<sup>3</sup>, n (%) | 17/604 (2.8) | 65/3480 (1.9) | 82/4084 (2.0) | 0.1 |
| Life satisfaction score<sup>4</sup>, mean (SD) | 18.1 (4.5) | 19.2 (4.0) | 19.0 (4.1) | <0.001 |
| Migrant, n (%) | 61/604 (10.1) | 125/3480 (3.6) | 186/4084 (4.6) | <0.001 |

<sup>1</sup> All risk factors were measured at wave 1, except for 'left school' which was defined as having left school at any wave.
<sup>2</sup> Two-tailed p-value of test for difference between those who did and did not self-report discrimination at waves 1 or 2. Chi-squared tests were used for binary variables and t-tests were used for continuous variables.
<sup>3</sup> Left school without obtaining a primary or secondary school qualification.
<sup>4</sup> Range: 5-25.
Note: Only twins with known zygosity (n=2,042 pairs) were included.

There was strong evidence for differences between groups with and without self-reported discrimination for mental disorder (*P*<0.001), sex (*P*=0.001), global life satisfaction score (*P*<0.001), and migrant status (*P*<0.001).

### Familial correlations

Sex-adjusted intra-class correlations (ICC) were 0.24 (95% Confidence Interval: 0.20–0.28) for mental disorder, 0.55 (95%CI: 0.51–0.58) for BMI, 0.49 (95%CI: 0.45–0.52) for current smoking status, 0.23 (95%CI: 0.19–0.27) for leaving school, and 0.36 (95%CI: 0.32–0.39) for global life satisfaction score. The sex-adjusted familial risk ratio for mental disorder was 6.54 (95%CI: 5.00–8.56, *P*<0.001).

### Within-individual models

Table 2 describes the within-individual unadjusted and adjusted associations between mental disorder and other risk factors with experiencing discrimination. In the adjusted within-individual model, having a mental disorder was positively associated with discrimination (adjusted OR=2.19, 95%CI: 1.42–3.39, *P*<0.001). There was also evidence of a negative association of male sex (aOR=0.62, 95%CI: 0.45–0.85, *P*=0.003) and life satisfaction (aOR=0.94, 95%CI: 0.91–0.98, *P*=0.001) with discrimination. In a separate model including migrant status (Table S3, Supplementary Material), this risk factor was strongly associated with discrimination in the unadjusted model (OR=4.97, 95%CI: 2.79–8.85, P<0.001), but this attenuated to the null in the adjusted model (aOR=2.86, 95%CI: 0.87–9.39, *P*=0.084).

**Table 2:**
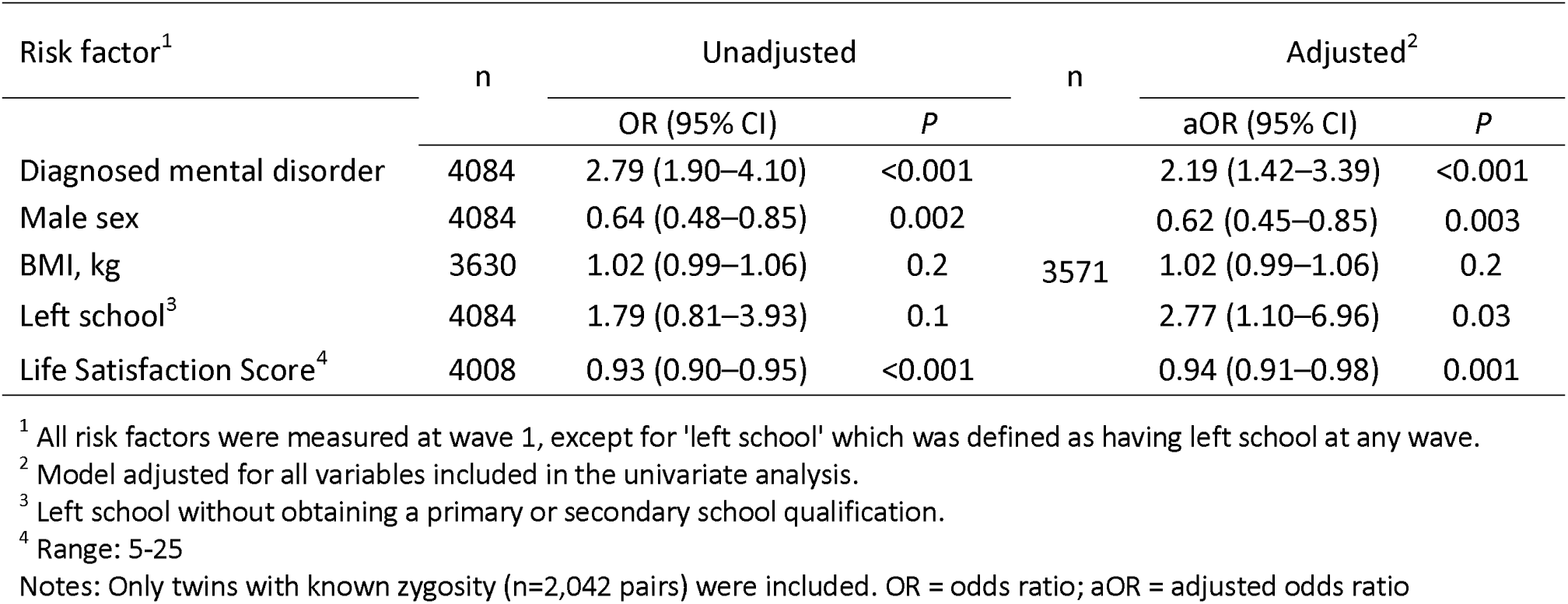
Within-individual unadjusted and adjusted associations between mental disorder and other risk factors with discrimination.

### Within-and-between pair models

Table 3 presents unadjusted and adjusted within-and-between pair associations between mental disorder and perceived discrimination. We did not find strong evidence between the within-pair difference of mental disorder or any covariates and discrimination in unadjusted or adjusted models. We found between-pair associations between diagnosed mental disorder (aOR=4.24, 95%CI: 2.17–8.29, *P*<0.001), male sex (aOR=0.63, 95%CI: 0.46–0.87, *P*=0.005), and life satisfaction (aOR=0.93, 95%CI: 0.89–0.97, *P*=0.001), after adjusting for covariates.

**Table 3:**
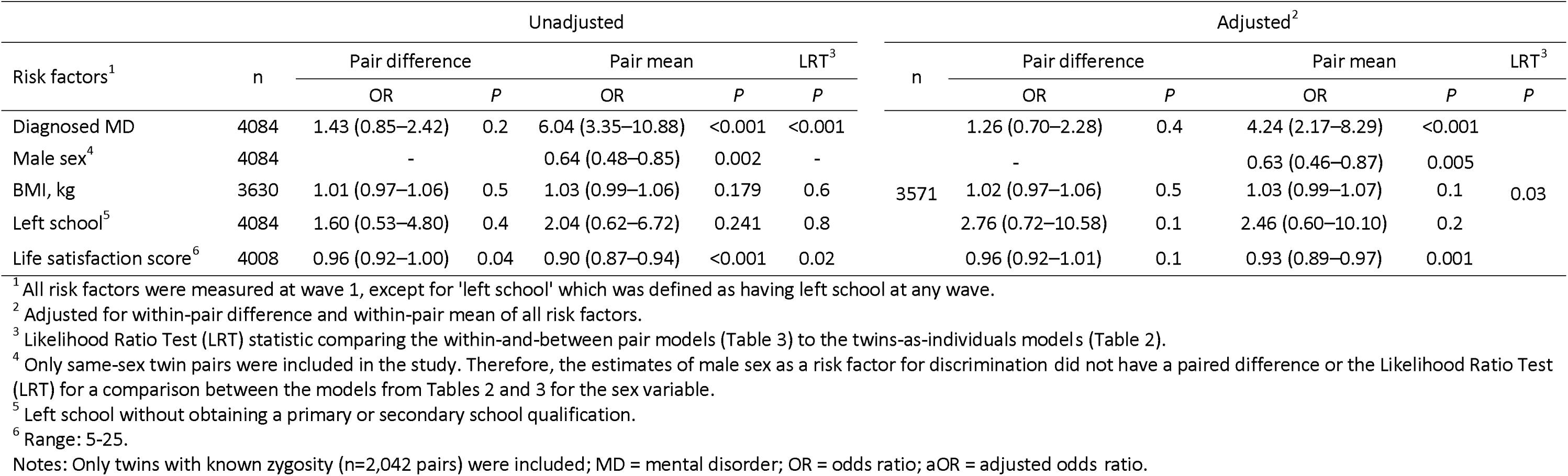
Within-and-between pair unadjusted and adjusted associations between mental disorder and other risk factors with exper ience of discrimination.

Likelihood ratio tests (LRTs) indicated that the within-and-between pair models had a better fit than within-individual models only for mental disorder (*P*<0.001) in the unadjusted models. The fully adjusted within-and-between model offered a marginal improvement over the within-individual model (*P*=0.03). Figure 1 presents a visual comparison between within-individual (Table 2), and within and between-pair estimates (Table 3) of the association between mental disorder and discrimination.

**Figure 1:**
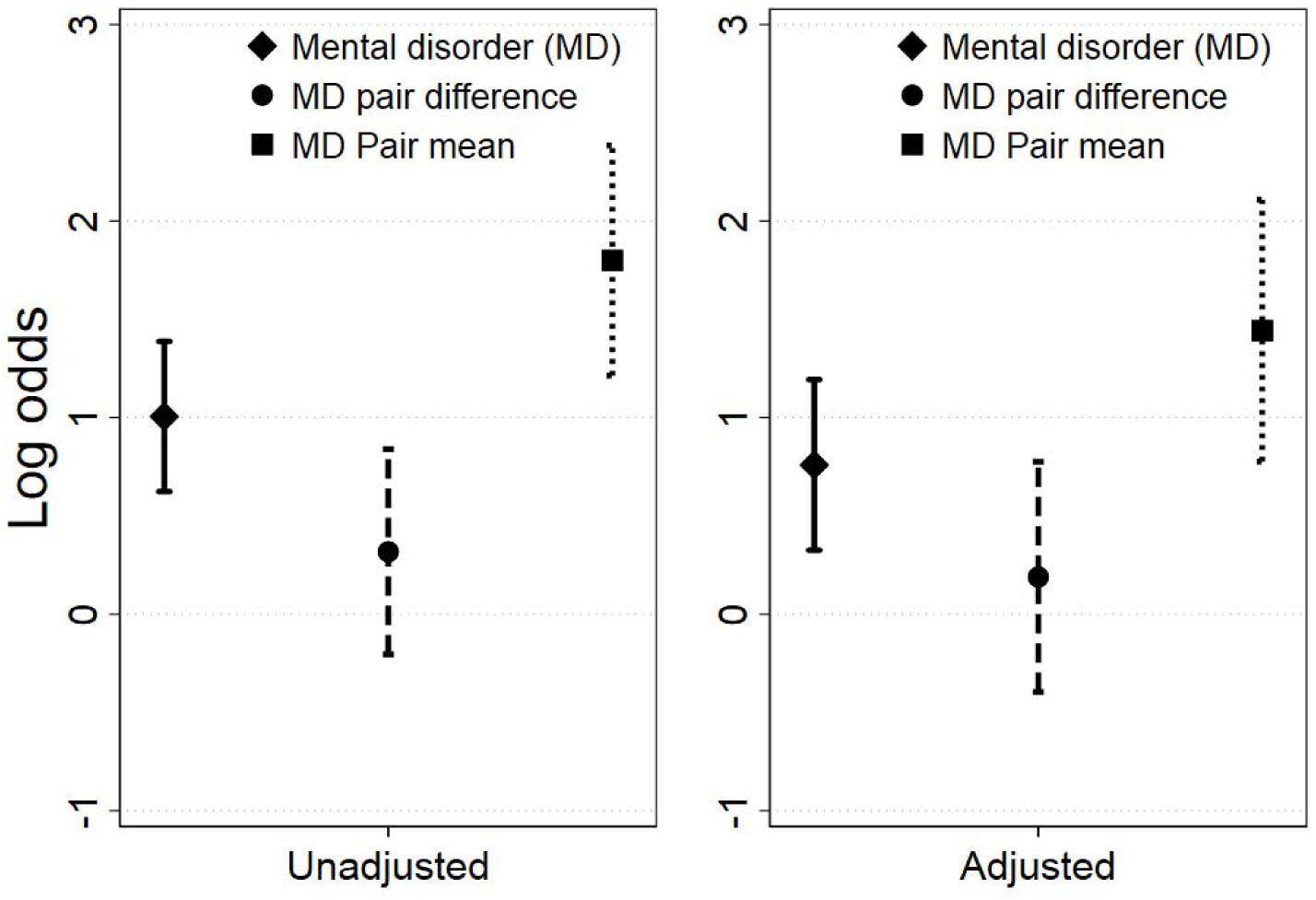
Log odds of unadjusted and adjusted associations between diagnosed mental disorder (within-individual model), and within-and-between pair difference models of diagnosed mental disorder with experience of discrimination at any wave, including 95% confidence intervals

### Sex differences

The sex differences analysis can be found in Table S5 (Supplementary Material). We found evidence of an interaction between male sex with the between-pair difference of mental disorder (*P*=0.003). In the analysis stratified by sex, we found differences between the between-pair difference of mental disorder for males (aOR=21.10, 95%CI: 5.82–76.56, *P*<0.001) and females (aOR=2.15, 95%CI: 0.96–4.77, *P*=0.06).

Our sensitivity analyses broadly supported our primary results. While the estimates in the sensitivity analyses were less precise due to studying only those who participated in both waves, we found no substantial differences in our main findings (Tables S1 and S2, Supplementary Material). Similarly, results from using mental illness (only anxiety and depression disorder) as an exposure (Table S4, Supplementary Material) did not produce considerably different results in the point estimates, although precision was again compromised.

## DISCUSSION

We investigated the association between mental disorder and perceived discrimination in young twin pairs in Germany, using both within-individual and within-and-between pair models to adjust for and assess the magnitude of familial confounding in this association. Our primary finding was that the within-individual association between having a mental disorder and experiencing discrimination, which we observed, attenuated to the null after adjusting for familial factors. This was evidenced by the presence of an association between the within-pair mean but not the within-pair difference in mental disorder with discrimination. We found formal evidence of familial confounding in the unadjusted association between mental disorder and discrimination, as indicated by the LRT favouring the within-and-between pair as opposed to the within-individual model, and in the fully adjusted model, although to a lesser extent. Further, our results provide no support for an association between mental disorder and discrimination when familial factors shared by twin pairs are adjusted for. Our findings indicate that the association between mental disorder and discrimination is almost fully explained by unmeasured familial factors.

Our results support the need for family-based mental health and discrimination interventions that incorporate family members of adolescents rather than exclusively focusing on individual behavioural factors. As mental disorder severity might be greater for individuals at higher genetic risk,^35^ incorporating a family history-informed approach to prevention and early intervention may improve both the efficiency and efficacy of these strategies.

Mental disorder and discrimination are systemic and pervasive problems with major social and economic repercussions for everyone. It is likely that larger, structural aspects of social determinants and how our society addresses mental health and discrimination play a role in this relationship. Societal efforts to address social inequalities that adversely affect families at higher risk of discrimination may be critical to achieving mental health equity. Thus, familial approaches to prevention should be complemented by universal interventions targeted at changing the determinants of stigma and discrimination more broadly.^36^ Nonetheless, mental health-related stigma and discrimination have been linked to a higher risk of unemployment, lower income, and higher healthcare costs.^37^ Therefore, it is critical to interrupt this cycle of social disadvantage and mental ill-health early in life, prior to its entrenchment throughout the life course.

Another explanation for the results we observed is that having a co-twin experiencing mental health-related or general discrimination might heighten one’s awareness of issues related to discrimination (due to mental health or other factors). This would result in more clustering of the reporting of such events in families and, therefore, reducing the number of pairs who are discordant for mental disorder or discrimination. The familial clustering in this association, as well as a potential bi-directional association, present important methodological challenges that might be potentially addressed using other novel twin and family study designs,^38^ and including data from future waves of the TwinLife Study in longitudinal analyses.

Our study also found that male twins from pairs where both were diagnosed with a mental disorder were at more than 21 (5.82–76.46) times higher odds of experiencing discrimination than male twins from unaffected pairs, compared to 2.15 (0.96–4.77) times higher odds for females, in the model adjusted for all covariates. This indicates that the familial confounding in the adjusted association between mental disorder and discrimination differs by sex and that the risk of discrimination as a function of a family history of mental disorder is greater for males than females. The observed sex differences also have ramifications for interventions. The finding that familial confounding is more present for young males than females may indicate that males are more susceptible to the impact of shared environmental factors on mental health and discrimination. Especially for males, family-based mental health interventions might better ameliorate the effects of discrimination. Understanding the familial determinants of adolescent and young adult mental health and discrimination should be both an area for future research and a public health priority.

Finally, we observed that being a migrant was associated with discrimination in the univariable within-individual model (OR=4.97, 95%CI: 2.79–8.85, *P*<0.001) but not in the multivariable model, indicating individual-level confounding. Nonetheless, it is clear that migrants were disproportionally exposed to discrimination events in this cohort and prevention efforts targeted at young migrant youth may be warranted.

Our study findings support the concept of ’intersectionality’, in which discrimination may be a result of multiple coalescing factors rather than just one cause,^12^ as it is likely determined by a complex network of interactions between genetic and environmental causes. This might also explain the well-known failure to reconcile estimates of (unmeasured) heritability of mental disorders and other health conditions found in twin studies with that observed in more recent genome-wide association studies with measured genetic variants.^39^ An approach to causal inference that considers genetic along with cultural and environmental differences within and between families within a wider set of social inequalities may yield better mental health and discrimination prevention strategies for adolescents and young adults. Studying any of these constructs while ignoring the other might represent a failure to achieve true equity and provide only temporary and insufficient answers to these major public health problems.

Our study had some limitations. The self-reported doctor’s diagnosis of mental disorder, as well as not having available data on the diagnoses of more stigmatised disorders (such as psychotic disorders), might have under-detected the true prevalence of mental disorder in our sample. Although any misclassification of this nature would result in a conservative measure of effect and therefore, this was unlikely to have considerably impaired this study. Our measure of self-reported discrimination was broad, and we did not have a sufficient sample size to investigate discrimination by specific causes. Recall bias might also have influenced our estimates.

The inability to establish a direct timeline between the onset (or diagnosis) of mental disorders, the other risk factors included in our models and discrimination as the outcome limited potential causal inference in our study. There is a possibility that some of the covariates, such as BMI and life satisfaction, might lie on the causal pathway between mental disorder and discrimination, and similarly for other risk factors studied in the adjusted models.

Our goal in this study was to assess potential familial confounding in the studied associations other than to confirm or falsify potentially causal associations. We were also less concerned with the potential of finding ’falsely significant’ within-individual or within-pair associations between mental disorder and discrimination and, therefore, did not adjust for multiple comparisons in our statistical analysis.^40^ Nonetheless, it is important to note that while a potentially causal association between mental disorder and discrimination appear to be unlikely – as evidenced by the ’null’ within-pair estimates we found in our results – this causal hypothesis can be formally tested using methods that allow for causal inference from twin and family data.

We intentionally grouped MZ and DZ pairs together in our regression analysis to increase the total sample size, precluding further disentangling genetic from shared environmental sources of confounding. Potential gene-environment correlation or interaction could not be tested in our models. Further research is needed to understand whether they may play a role in the association between mental disorder and discrimination.

While the within-family study design here employed is an efficient tool to investigate associations holding familial (including genetic) factors constant, it can also be less successful in fully considering differences between genetically and culturally diverse groups in our society. Importantly, the proportion of migrants was lower, and levels of education and income were slightly higher in the TwinLife study compared to the general German population.^22^ Even considering that TwinLife used a population-based recruitment strategy, this indicates that our findings might not be directly generalisable to all contexts.

## Conclusions

Our study is the first to provide evidence that the association between having a mental disorder and experiencing discrimination is confounded by familial factors shared by twin pairs, such as genetic and shared environmental factors, and household social determinants of health. Incorporating family members in interventions targeted at ameliorating mental ill-health and experiences of discrimination among adolescents may improve efficacy, especially for males.

## Declaration of Interest

GN-H is a deputy editor of BJPsych but played no part in the editorial process or decision making of this manuscript. All other authors have no interest to declare.

## Funding

The TwinLife Project is funded by the German Research Foundation (grant number 220286500). LCF is funded by a Suicide Prevention Australia Postgraduate Fellowship. JTY receives salary and research support from a National Health and Medical Research Council Investigator Grant (GNT1178027).

## Data Availability

Data is available from the data custodian (Gesis - gesis.org) upon reasonable request.

http://gesis.org

## Acknowledgements

We acknowledge all participants from the TwinLife study for their invaluable contribution.

## Author contribution

LCF designed the study and the analytical strategy, conducted the statistical analysis, interpreted the findings and drafted the manuscript. JTY designed the study and the analytical strategy, interpreted the findings and revised the manuscript. GA, PK and JLH designed the analytical strategy, interpreted the findings and revised the manuscript. FMS and EH facilitated the access and the understanding of the TwinLife study data, interpreted the findings and revised the manuscript. GNH and JF interpreted the findings and revised the manuscript. All authors approved the final version of this manuscript.

## Notes

### Competing Interest Statement

The authors have declared no competing interest.

