## Supplementary Material for "Mental disorders and discrimination: a prospective cohort study of young twin pairs in Germany"

| Table S1: Unadjusted and adjusted associations between mental disorder and other risk factors at wave 1 and discrimination (at wave 2) | | | | | |
| --- | --- | --- | --- | --- | --- |
|  |  | Unadjusted | | Adjusted | |
|  |  | OR | *p* | aOR | *P* |
| Diagnosed mental disorder |  | 2.96 (1.81–4.85) | <0.001 | 2.73 (1.54–4.82) | 0.001 |
| Male sex |  | 0.51 (0.34–0.75) | 0.001 | 0.50 (0.33–0.77) | 0.001 |
| BMI, kg |  | 1.01 (0.96–1.06) | 0.7 | 1.01 (0.96–1.06) | 0.6 |
| Left school |  | 2.27 (0.73–7.05) | 0.2 | 2.92 (0.83–10.30) | 0.1 |
| Satisfaction score |  | 0.93 (0.89–0.97) | <0.001 | 0.96 (0.92–1.00) | 0.06 |
| Note: this analysis was restricted to participants who participated in both waves of the study. See methods for details. | | | | | |

| Table S2: Unadjusted and adjusted associations between risk factors and discrimination (at both waves) | | | | | |
| --- | --- | --- | --- | --- | --- |
|  |  | Unadjusted | | Adjusted | |
|  |  | OR | *p* | aOR | *P* |
| Diagnosed mental disorder |  | 3.81 (1.81–8.01) | <0.001 | 3.74 (1.57–8.92) | 0.003 |
| Male sex |  | 0.55 (0.30–1.02) | 0.06 | 0.52 (0.26–1.04) | 0.07 |
| BMI, kg |  | 1.01 (0.94–1.09) | 0.7 | 1.02 (0.95–1.10) | 0.6 |
| Left school |  | 1.59 (0.26–9.56) | 0.6 | 1.93 (0.26–15.14) | 0.5 |
| Satisfaction score |  | 0.93 (0.87–0.99) | 0.02 | 0.96 (0.89–1.03) | 0.3 |
| Note: this analysis was restricted to participants who participated in both waves included in this study. See methods for details. | | | | | |

| Table S3: Unadjusted and adjusted associations between mental disorder and other risk factors with discrimination (at any wave), including migrant variable | | | | | |
| --- | --- | --- | --- | --- | --- |
|  |  | Unadjusted | | Adjusted | |
|  |  | OR | *p* | aOR | *P* |
| Diagnosed mental disorder |  | 2.79 (1.90–4.10) | <0.001 | 3.91 (1.74–8.78) | 0.001 |
| Male sex |  | 0.64 (0.48–0.85) | 0.002 | 0.48 (0.24–0.96) | 0.04 |
| BMI, kg |  | 1.02 (0.99–1.06) | 0.2 | 1.00 (0.94–1.08) | 0.9 |
| Left school |  | 1.79 (0.81–3.93) | 0.1 | 1.51 (0.22–10.56) | 0.7 |
| Satisfaction score |  | 0.93 (0.90–0.95) | <0.001 | 0.96 (0.90–1.03) | 0.2 |
| Migrant |  | 4.97 (2.79–8.85) | <0.001 | 2.86 (0.87–0.06) | 0.08 |

| Table S4: Unadjusted and adjusted associations between mental illness and other risk factors with discrimination (at any wave) | | | | | |
| --- | --- | --- | --- | --- | --- |
|  |  | Unadjusted | | Adjusted | |
|  |  | aOR | *p* | aOR | *p* |
| Diagnosed mental illness |  | 2.35 (1.44–3.83) | 0.001 | 2.63 (0.93–7.40) | 0.07 |
| Male sex |  | 0.64 (0.48–0.85) | 0.002 | 0.48 (0.24–0.95) | 0.03 |
| BMI, kg |  | 1.02 (0.99–1.06) | 0.2 | 1.00 (0.93–1.08) | 1.0 |
| Left school |  | 1.79 (0.81–3.93) | 0.1 | 1.71 (0.25–11.61) | 0.6 |
| Satisfaction score |  | 0.93 (0.90–0.95) | <0.001 | 0.95 (0.88–1.02) | 0.1 |
| Note: Mental illness was defined as diagnosed anxiety or depression. | | | | |  |

| Table S5: Adjusted associations between within-pair difference and pair within-pair mean of risk factors (at wave 1) with experience of discrimination (at any wave) from within-and-between pair models, separately for males and females with an interactive term | | | | | | | | | | | |  |
| --- | --- | --- | --- | --- | --- | --- | --- | --- | --- | --- | --- | --- |
|  | Males | | | | |  | | Females | | | | Interaction with sex^1^ |
| Risk factors | Within-pair difference | | Between-pair difference | | |  | | Within-pair difference | | Between-pair difference | |  |
|  | aOR | *P* | aOR | *P* |  | | aOR | | *P* | aOR | *P* | P |
| Diagnosed mental disorder | 1.23 (0.45–3.33) | 0.7 | 21.10 (5.82–76.56) | <0.001 |  | | 1.25 (0.60–2.58) | | 0.6 | 2.15 (0.96–4.77) | 0.06 | 0.003 |
| BMI, kg | 1.01 (0.93–1.09) | 0.9 | 1.05 (0.98–1.13) | 0.1 |  | | 1.02 (0.97–1.07) | | 0.5 | 1.02 (0.98–1.07) | 0.3 | 0.4 |
| Left school | 10.41 (1.14–94.72) | 0.04 | 1.07 (0.12–9.77) | 1.0 |  | | 1.03 (0.15–7.03) | | 1.0 | 3.77 (0.51–27.99) | 0.2 | 0.6 |
| Satisfaction score (X-Y) | 0.94 (0.87–1.01) | 0.09 | 0.93 (0.87–1.00) | 0.04 |  | | 0.98 (0.92–1.03) | | 0.4 | 0.93 (0.88–0.98) | 0.009 | 0.7 |
| ^1^ P-values of interactive term for sex and the between-pair difference of each risk factor. These estimates were obtained from a different model to those for males and females which were stratified and not adjusted by sex. | | | | | | | | | | | | |
